# Predicting the evolution of SARS-Covid-2 in Portugal using an adapted SIR Model previously used in South Korea for the MERS outbreak

**DOI:** 10.1101/2020.03.18.20038612

**Authors:** Pedro Teles

**Affiliations:** Departamento de Física e Astronomia, Faculdade de Ciências da Universidade do Porto, Rua do Campo Alegre s/n, 4169-007 Porto

**Keywords:** covid-19, epidemiological assessment, SIR model

## Abstract

The covid-19 has spread very quickly worldwide, leading the World Health Organization (WHO) to declare a state of pandemic. Moreover, the WHO has announced that the European continent is now the main centre of the pandemic.

One of the questions many governments are asking is how the spread is going to evolve in time. In this study, an adapted SIR model previously used in South Korea to model the MERS outbreak was applied to estimate the evolution of the curve of active cases in the case of the Portuguese situation. As some of the parameters were unknown, and the data for Portugal is still scarce, given that the outbreak started later (first case on the 2nd of March) I used Italian data (first reported case in Italy on the 31st of January) to predict them. I then construct five different scenarios for the evolution of covid-19 in Portugal, considering both the effectiveness of the mitigation measurements implemented by the government, and the self-protective measures taken by the population, as explained in the South Korean model.

In the out of control scenario, the number of active cases could reach as much as 40,000 people by the beginning of April. In the best-case-scenario considered, the active cases could reach circa 7,000 people. The actual figure probably lies between the interval (7,000-13,000) and the peak will be reached between 9th and the 20th of April 2020.

Without control and self-protective measures, this model predicts that the figures of active cases of SARS-covid-2 would reach a staggering 40,000 people It shows the importance of control and self-protecting measure to bring down the number of affected people by following the recommendations of the WHO and health authorities. With the appropriate measures, this number can be brought down to 7,000-13,000 people

## 1 Introduction

There is already abundant information on covid-19[1-5]. In the most severe cases, covid-19 can lead to the development of acute respiratory distress syndrome (ARDS) causing respiratory failure, septic shock, multiorgan failure, and even death [6]. Studies suggest that the case fatality rate of the virus is of about 3.5% in mainland China [7]. However, this value seems to be much higher in Italy [8], suggesting its strong dependence on demographics [9]. The WHO declared Europe as the new epicentre of the disease on the 13th of March of 2020[10]. The rapid growth of the number of active cases presenting severe symptoms has saturated the health services in most countries in the continent, especially in Italy [11]. Governments throughout Europe have implemented severe measures to prevent and mitigate the spread of the virus. Yet, as of the 20th of March there were as many as 129,811 confirmed cases on the European continent, resulting in 6,069 deaths[2, 3]. In Portugal, the two first confirmed cases in the Portuguese territory were announced on the 2nd of March,.As of the 20^th^of March, there were 1,020 confirmed cases, 6 deaths, and 5 recovered [12]. Also, the country is on lockdown since the 13th of March 2020, and the Portuguese Parliament declared a State of Emergency on the 18th of March 2020. Many models have been used to study the spread of covid-19 [13, 14], In this study, a model of the SIR type used in South-Korea to model the evolution of the active cases of the MERS outbreak in the country back in 2015 [15] was adapted. In order to fit the parameters, and given that there is still very little data in the case of Portugal, the data of Italy was used. This allowed to fit the curve of current active cases in Portugal with a model, which is then used to predict the future number of cases, in five different scenarios.

## 2 Methodology

The model used is described in Xia et al[15], which can be understood by the flow diagram shown in figure 1 (which was taken from the paper, page 4). In this model, S corresponds to the number of susceptible individuals; E, the number of exposed individuals; A, the number of asymptomatic infected cases; I, the total number of mild-to-severe infected patients. H, the number of hospitalized cases; R, the number of removed cases, and finally N, the total population of Portugal.

**Figure 1:**
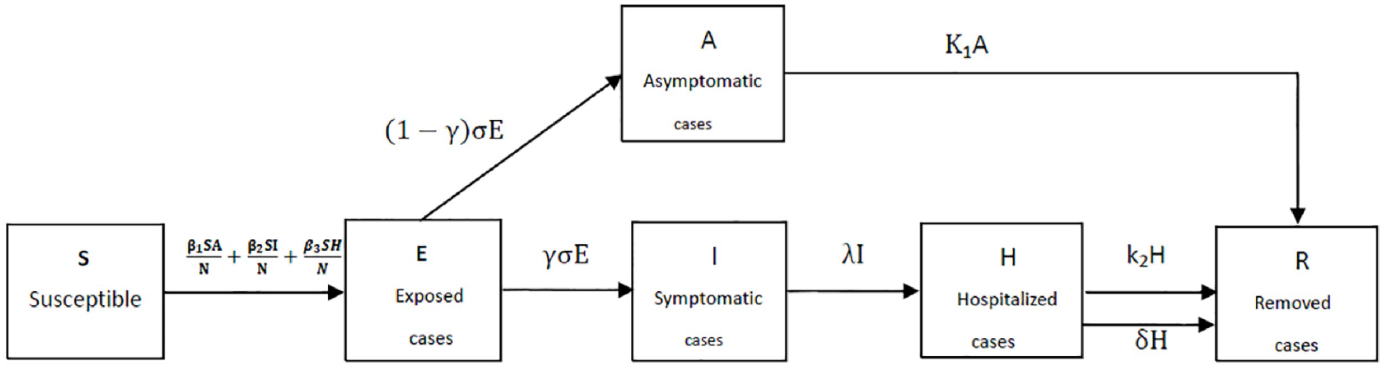
Flow chart of the SIR-type model used in this work, taken from [15]

*β*_1_ is the transmission coefficient of the asymptomatic infected cases, *β*_2_ is the transmission coefficient of the symptomatic infected cases (mild infected person and severe patients) to the susceptible, *β*_3_ is the transmission coefficient of the hospitalized cases to the susceptible, *σ*^−1^ is the mean incubation period, *λ*^−1^ is the mean time between symptom onset to hospitalization, k_1_^-1^ is the mean infectious period of asymptomatic infected person for survivors, k_2_^-1^ is the mean duration for hospitalized cases for survivors, *δ*^−1^ is the mean time from hospitalization to death, *γ* is the clinical outbreak rate in all the infected cases. The time unit is 1 day.

The set of differential equations can then be written as:

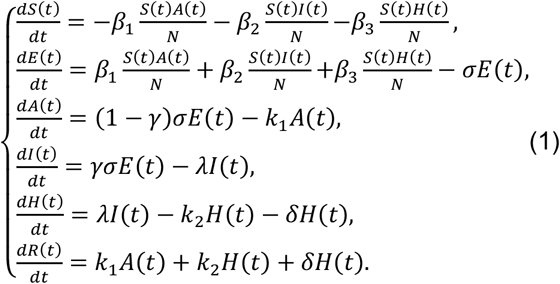

The model was further adapted. the only parameters that were considered “known” were the mean incubation time, which was taken as *σ*^−1^=5.1 days from the literature (here using the median as equal to the mean) [16], and the mean infectious period of asymptomatic people, in which a conservative estimate of k_1_^-1^=14 days was used. Furthermore, *β*_1_ = *β*_2_ = *β*_3_= *β* was taken as a first approximation, meaning that the mean transmission coefficient is the same independently of type and setting of the transmission. Finally, the epidemic starts with only one person infected (I_0_=1), being the rest of the values 0, except for *S*_0_ which is simply N-1. See table 1 for a breakdown of the different parameters of this model.

**Table 1:** The different parameters of the model described by Eq. 1

| parameter | Xia et al values [15] | this work |
| --- | --- | --- |
| $\sigma^{-1}$ | 5.2 (3~8) (days) | 5.1 (days) |
| $\lambda^{-1}$ | 5(3~7) (days) | To be fitted |
| $\delta^{-1}$ | 15.16(0~42) (days) | To be fitted |
| $k_1^{-1}$ | 5 (days) | |
| $k_2^{-1}$ | 7 (days) | |
|  |  | To be fitted |
| $\beta_1$ | 0.8756(0.853~0.9324) (*days*<sup>-1</sup>) | To be fitted |
| $\beta_2$ | 0.7833(0.5925~0.8592) (*days*<sup>-1</sup>) | To be fitted |
| $\beta_3$ | 0.4568(0.3839~0.6751) (*days*<sup>-1</sup>) | To be fitted |
| $\gamma$ | 0.0348 (0.0285~0.353) (*days*<sup>-1</sup>) | To be fitted |
| N | 49,520,000 | 10,290,000 |
| $S_0$ | 49,519,960 | 10,289,999 |
| $E_0$ | 16~32 | 0 |
| $A_0$ | 16~36 | 0 |
| $I_0$ | 1 | 1 |
| $H_0$ | 0 | 0 |
| $R_0$ | 0 | 0 |

The solving of the system of differential equations was performed using the Mathematica code [17], using the function “NonLinearModelFit”[18].

In order to determine the parameters *λ* and *δ*, the number of deaths, as reported by the Italian ministry of Health and available online, was used [19]. For the estimation of the value of *k*_2_, the number of recovered was used. All data was taken until the 19th of March 2020.

After determining the values for these parameters, they were used to fit the curve of infected cases for Portugal, where the values of *β*_1_ = *β*_2_ = *β*_3_ = *β* and *γ* were fitted to obtain a model. This model was then used to estimate the evolution of active cases in Portugal, according to five different scenarios:

- Out-of-control scenario (nothing done, the virus is free to spread (out-ofcontrol scenario);
- Scenario in which the government takes severe mitigating measures and the population adheres to self-protecting measures equivalent to the original model (scenario 1)
- Scenario where government measures are 50% as effective as those in South Korea, and self-protective measures reduce the transmission rate by 50% (scenario 2)
- Scenario where government measures are 50% as effective as those in South Korea, and self-protective measures reduce the transmission rate by 70% (scenario 3)
- Scenario where government measures are 50% as effective as those in South Korea, and self-protective measures reduce the transmission rate by 80% (scenario 4)

## 3 Results

### 3.1 Fitting of parameters *λ* and *δ* (Italy)

The number of deaths in Italy was taken from [19], and the set of differential equations described in eq. 1 fitted to obtain the best possible fit for parameters *λ* and *δ*. An *R*^2^ value of 0.99255 was obtained with the fit. The rest of the parameters are presented in table 2. Results are shown in figure 2.

**Table 2:** fitted parameters *λ* and *δ* as obtained using “NonLinearCurveFit” in Mathematica

| parameter | estimate | standard error | t-statistic | p-value |
| --- | --- | --- | --- | --- |
| $\lambda$ | 0.2548 | 0.0002 | 879 | $9.83 \times 10^{-70}$ |
| $\delta$ | 0.0168 | 0.0002 | 73 | $3.02 \times 10^{-36}$ |

**Figure 2:**
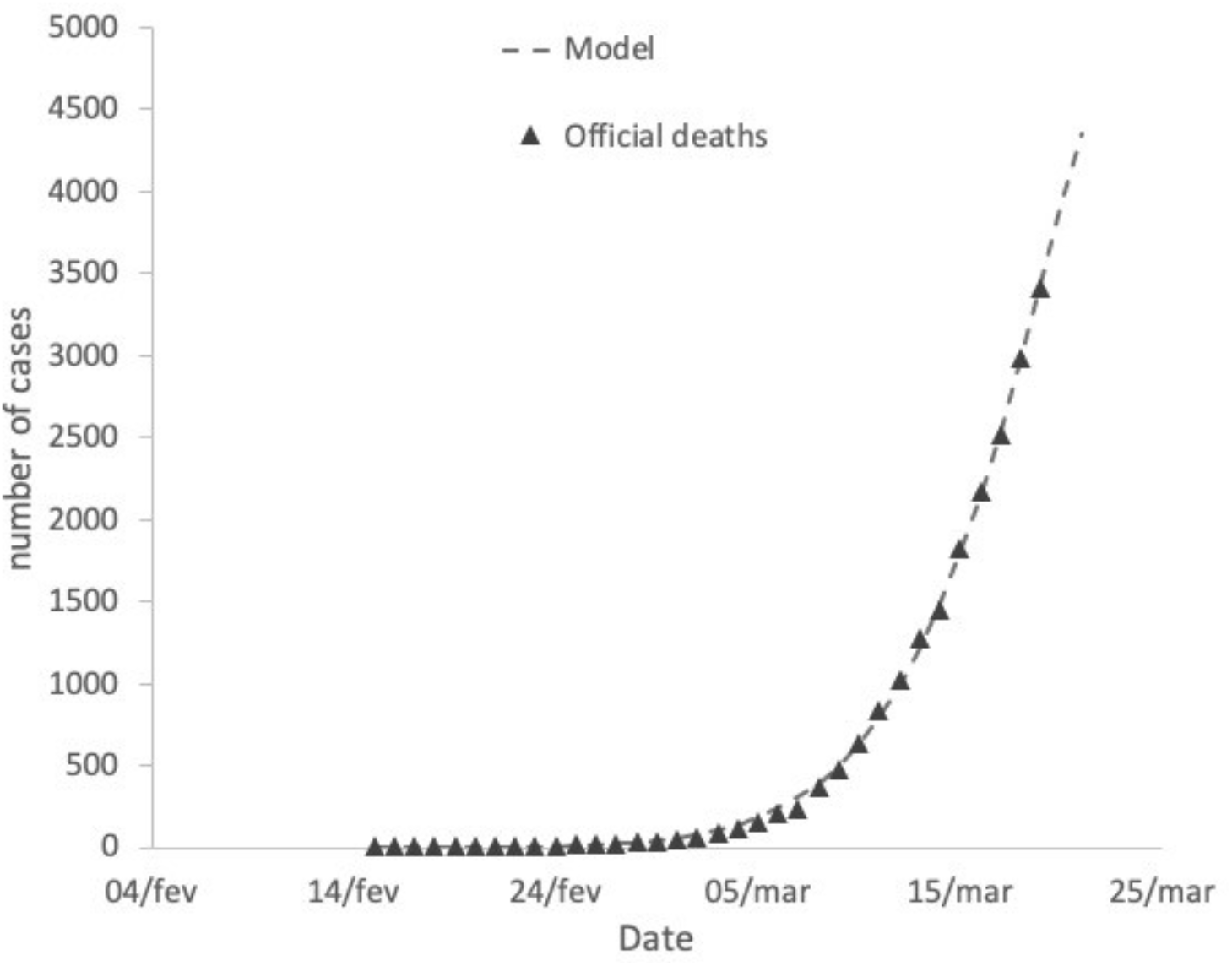
Graphical representation of the fitted model to the Italian government official death toll

### 3.2 Fitting of parameter *k*_2_ (Italy)

Now, using the values for *λ* and *δ* found in 3.1, parameter *k*_2_ was obtained with a new fit of Eq. 1, but now to the number of recovered cases in Italy, which again was taken from [19]. The best possible fit for *k*_2_ gave an *R*^2^ value of 0.86178.

The rest of the parameters are presented in table 3.

**Table 3:** Fitted parameter *k*_2_ as obtained using “NonLinearCurveFit” in Mathematica

| parameter | estimate | standard error | t-statistic | p-value |
| --- | --- | --- | --- | --- |
| $k_2$ | 0.0875 | 0.012 | 7.32 | $3.09 \times 10^{-9}$ |

As can be seen, in the case of recovered cases, the fit has not the same quality as the fit in 3.1. This is further evidenced by figure 3, where a graphical representation of the fit is shown.

**Figure 3:**
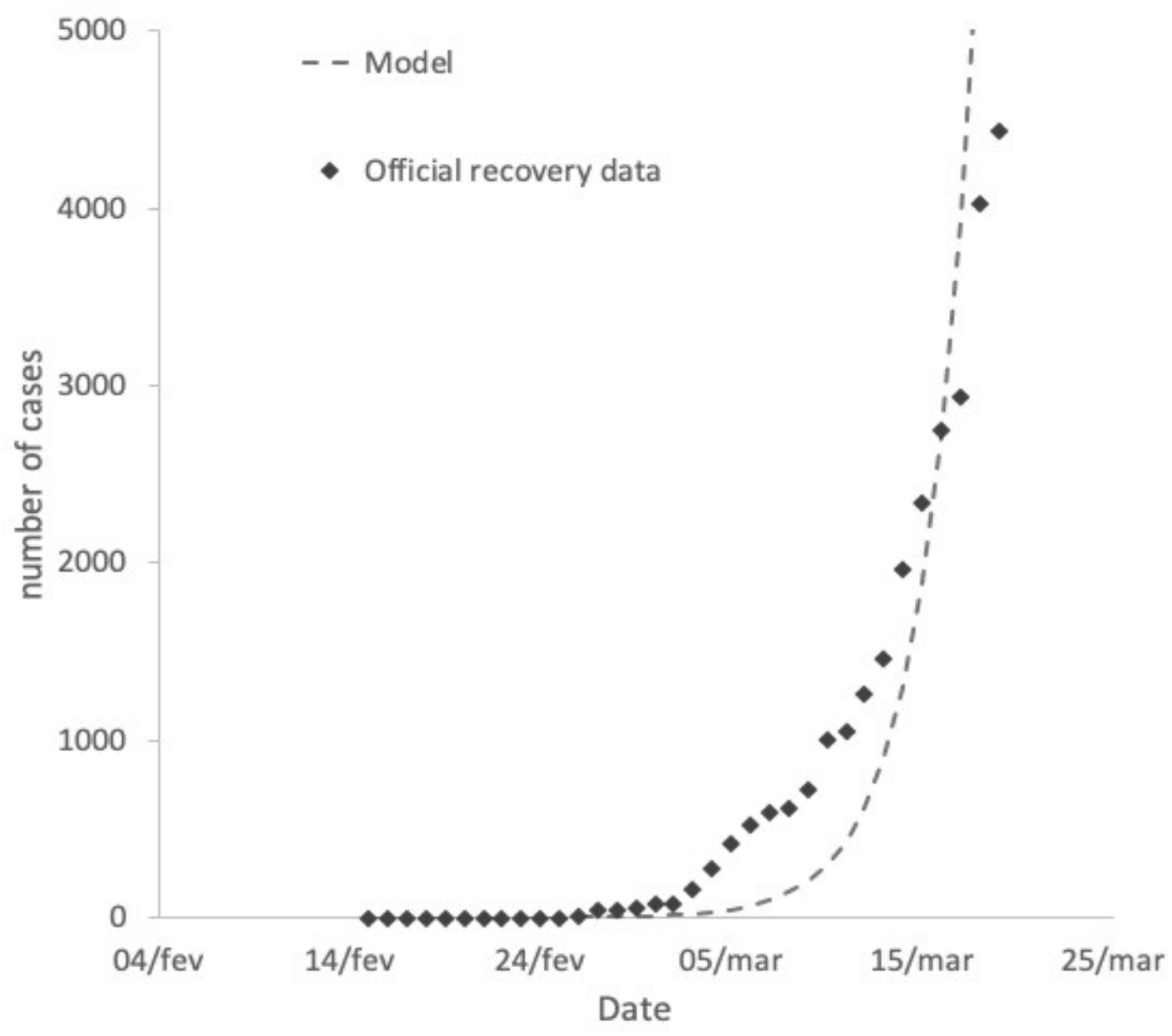
Graphical representation of the fitted model to the Italian government official recoveries.

### 3.3 Fitting of parameters *β* and *γ* (Portugal)

The parameters needed to fit Eq. 1 to the number of Portuguese active cases until the 19th of March, in order to determine the value of *β* and *γ*, were obtained in 3.1, and 3.2. The number of Portuguese active cases was taken from [20], which is updated daily.

The values previously obtained of parameters *λ, δ*, and *k*_2_ were used. As explained previously, because the fitting was difficult to obtain, as a first approximation, *β*_1_ = *β*_2_ = *β*_3_ = *β* was considered. The obtained values for *β* and *γ* are given in table 5.

**Table 4:** Fitted parameters *λ* and *δ* as obtained using “NonLinearCurveFit” in Mathematica

| parameter | estimate | standard error | t-statistic | p-value |
| --- | --- | --- | --- | --- |
| $\beta$ | 1.16 | 0.033 | 34.997 | $1.51 \times 10^{-16}$ |
| $\gamma$ | 0.0158 | 0.0034 | 4.634 | $2.74 \times 10^{-5}$ |

**Table 5:** The obtained parameters of the model described in the previous sections

| parameter | this work ($days^{-1}$) |
| --- | --- |
| $\sigma$ | 5.1(fixed) |
| $\lambda$ | $0.2548 \pm 0.0002$ |
| $\delta$ | $0.0168 \pm 0.0002$ |
| $k_1$ | 0.0714 (fixed) |
| $k_2$ | $0.0875 \pm 0.012$ |
| $\beta$ | $1.16 \pm 0.033$ |
| $\gamma$ | $0.0158 \pm 0.034$ (Portugal)* |

A graphical depiction of the fitted model is given in figure 4.

**Figure 4:**
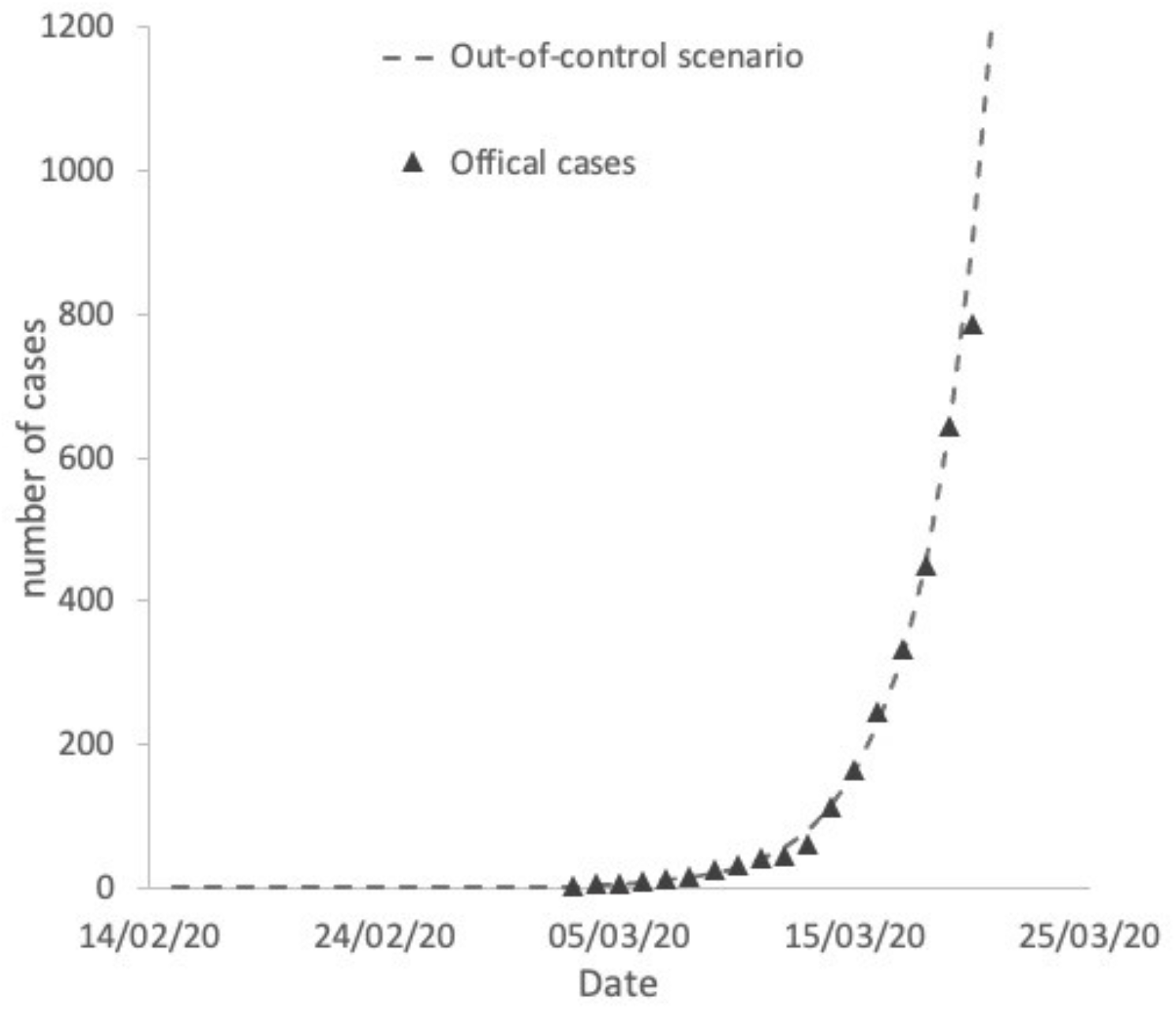
Graphical representation of the fitted model to the Portuguese government official active cases.

In this scenario, the out-of-control scenario, a peak of 40,000 infected would be reached at around the 5th of April 2020.

### 3.4 Discussion of the obtained values for *λ, δ, k*_2_, *β*, and *γ*

In order to obtain the parameters for *λ, δ*, and *k*_2_, an ansatz value for *γ* had to be given. In this model, *γ* is originally defined as “the clinical outbreak rate in all the infected cases. In the cases of the fits of *λ, δ*, and *k*_2_, the ansatz value used for *γ* had a very strong impact in the results of the other parameters. This could be due to two things: the huge uncertainty there is at the moment for the appropriate incubation time of this virus, which some authors claim can be of up to 27 days [21-22], and the uncertainty in how many in the symptomatic and asymptomatic cases are actually being tested. The model assumes that only those symptomatic are being tested which seems like a good approximation, but it comes with a lot of uncertainty.

The obtained value for *δ* = 0.0168 is 24% the value obtained by Xia et al[15] of 0.0660, which is just about the ratio in case fatality rates between the SARS-covid-2 (7.2% currently for Italy), and MERS (estimated at 34%) This seems reasonable, although the case fatality rate of the SARS-covid-2 is still difficult to ascertain.

Finally, from the obtained values it is also possible to determine a value for the basic reproduction number *R*_0_, which here I take as being simply the value R_0_=2*β/(k_1_+k_2_)=14.5 which is extremely high, and demonstrates how strong the capacity for spreading of this particular virus is. In table 5, the final values of the obtained parameters are given.

## 4 Five different scenarios for the evolution of SARS-covid-2 in portugal

The paper by Xia *et al*[15] further presents an extra 7 parameters to take into account control measures in time evolution of the epidemic. These parameters are meant to take into account two things:

- Isolation and monitoring measures taken by the government (parameters *d*_1_, *d*_2_, *d*_3_, and *d*_4_);
- Self-protection measures taken by the population (parameters *l*_1_, *l*_2_, and *l*_3_). The way these parameters are introduced in the model are as follows:

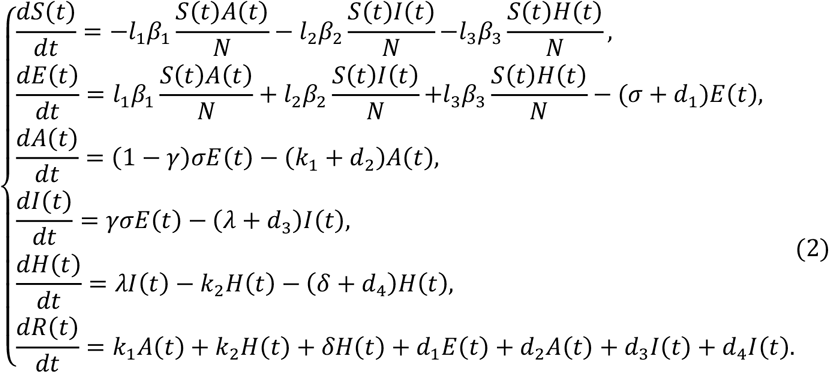

The *l*_1_, *l*_2_, and *l*_3_ parameters are basically multiplication factors for the transmission parameter *β*, so in this model we only consider one value for it as in the previous sections. The *d*_1_, *d*_2_, *d*_3_, and *d*_4_ parameters are connected to implemented government measures. The values provided by Xia *et al*[15], when applied to the Portuguese case, seem to provide a very unrealistic scenario. As such, four different scenarios in addition to the “out-of-control” scenario described in the previous section, were devised as explained in section 2. This is the out-of-control scenario, scenario 1, scenario 2, scenario 3, and scenario 4.

An educated guess were made that protective measures would diminish the transmission rate by 50%. For the government related measures the same value as the ones from Xia *et al*[15] were used. In the second scenario the 50% reduction is kept in the transmission rate but the efforts of the government are reduced by one half. In the third and fourth scenarios 70% and 80% of the transmission rateare used respectively.

The values for these scenarios are given in table 6.

**Table 6:**
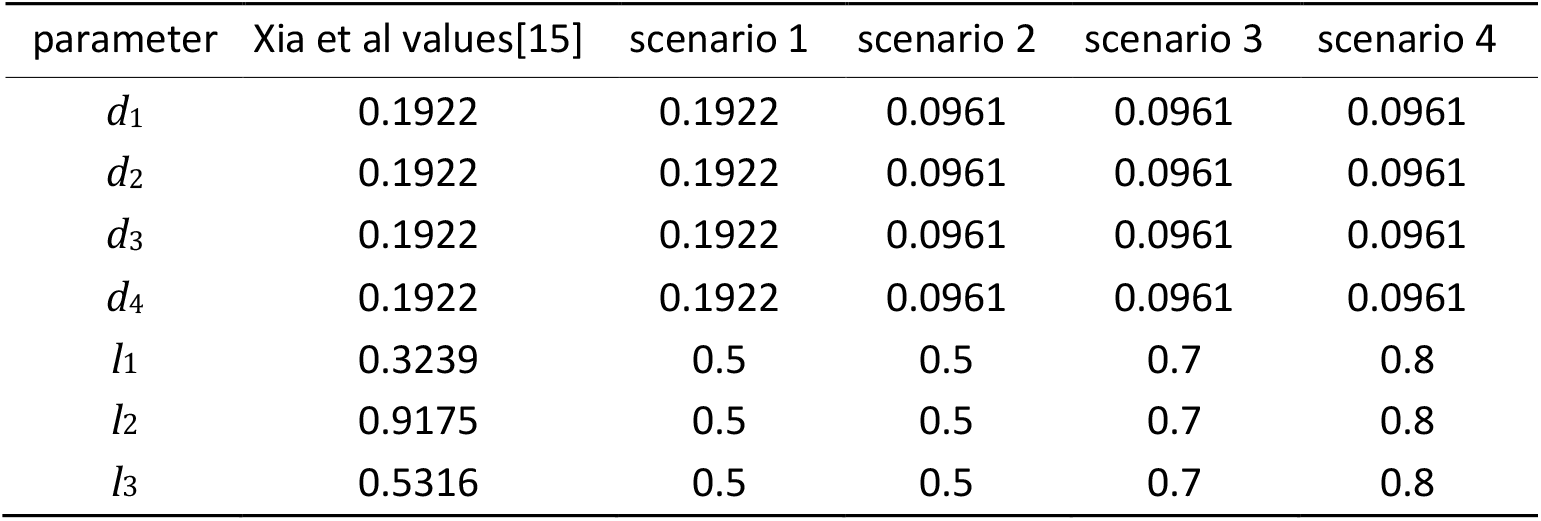
Control parameters of the model described by Eq. 2

The day considered for the initiation of these measures is day 33 corresponding to the 18th of March 2020 which is 5 days after the government implemented the measures, so giving the virus another cycle of infection before the measures take effect. Results are shown in figure 5.

**Figure 5:**
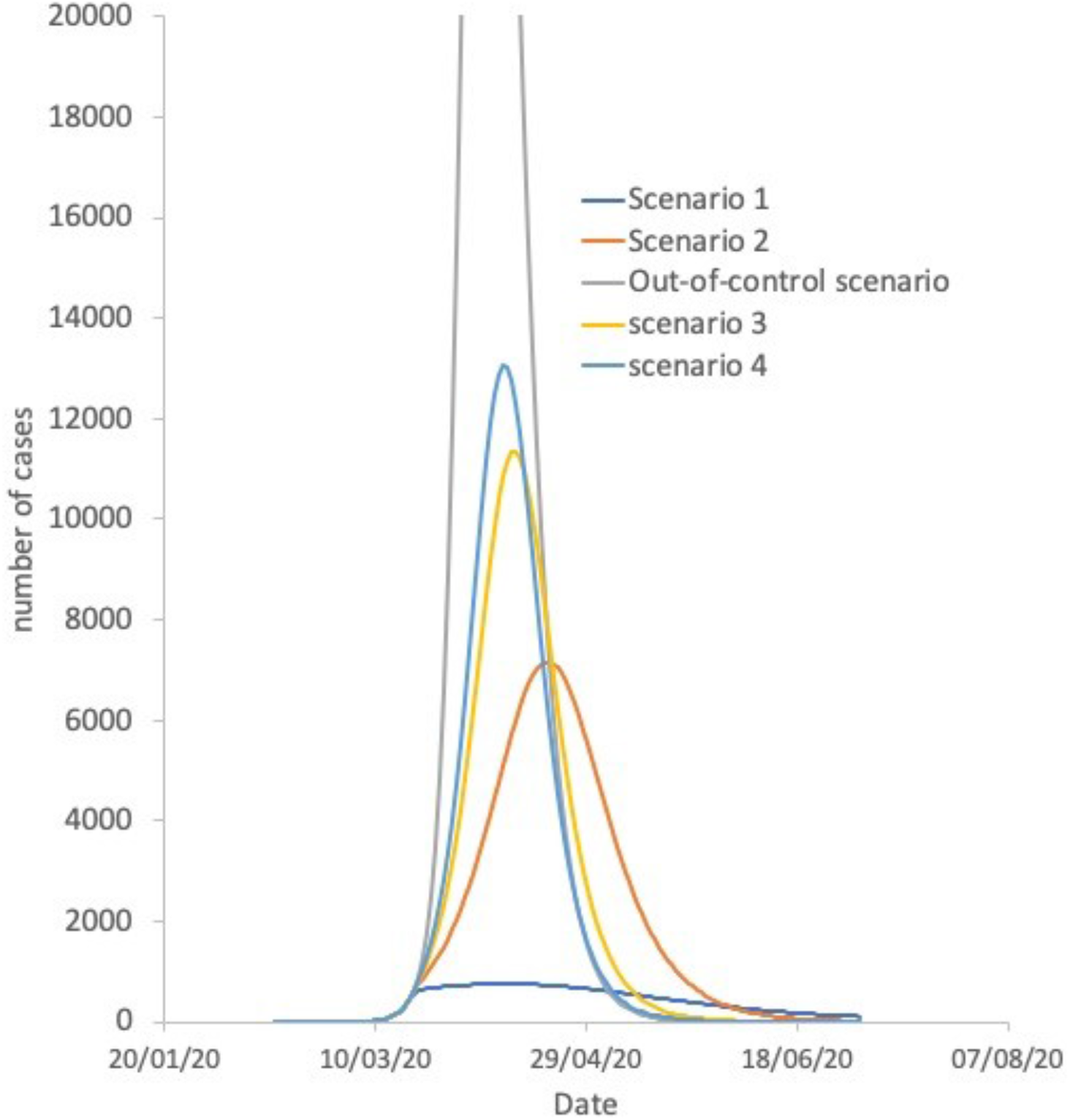
Graphical representation of the different scenarios considered in this study.

## 5 Conclusions

Results show that measures can be effective in “flattening the curve”. In the first scenario (unrealistic) the effectiveness of the control measures is maximum and just about 800 cases would have been reached. This has proven to be an unrealistic scenario given that as of today there are 1600 active cases in the country. In scenario 2, the maximum is shifted to the 20th of April, reaching a maximum of 7,000 active cases in the country. In this scenario, the transmission rate has been cut to a half, but the government’s measures effectiveness has been cut to a half of the value from the original model used in Korea. In scenarios 3 and 4, the measures of the government’s effectiveness are kept at the same value, but the transmission rate is only cut to 70% and 80%, respectively. In scenario 3 the number of infected woulf reach a peak of about 11,000 people, on the 12th of April, and finally in scenario 4, the peak would be reached at 13,000 cases, on the 9th of April. In the out-of-control scenario, the peak would be reached at about 40,000 people on the 5th of April.

## 6 Limitations of this study and scope of application

The current work makes a series of assumptions which may or may not be proven correct in the future, including the incubation period of the virus, which is still not well-known. This proved to be a major source of uncertainty to fit the results, because it had a great influence in the other parameters, especially parameter *γ*. Instead this parameter seems to behave in this study as an indicator of the huge uncertainty of the value for the incubation period, and testing capabilities. This model takes the values of active cases and compares them with official confirmed numbers, which not only may be smaller than the actual cases, as are released at an administratively defined time, which does not correspond to something realistic. And although testing is probably mostly done to the symptomatic cases described the fact is that many asymptomatic cases can have also been tested and therefore constitute another source of uncertainty.

The maximum number of active cases in Portugal will likely be in the interval 7,000-13,000 between the 9th-20th of April, if the government and the people of Portugal manage to keep implementing self-protective and control measures.

## Data Availability

All of it available publically.

